# A spatio-temporal study of state-wide case-fatality risks during the first wave of the COVID-19 pandemic in Mexico

**DOI:** 10.1101/2021.07.22.21260793

**Authors:** Ricardo Ramírez-Aldana, Juan Carlos Gomez-Verjan, Omar Yaxmehen Bello-Chavolla, Lizbeth Naranjo

## Abstract

We study case-fatality risks (risks of dying in sick individuals) corresponding to the first wave of the COVID-19 pandemic in Mexico. Spatio-temporal analysis by state were performed, mainly from April to September 2020, including descriptive analyses through mapping and time series representations, and the fit of linear mixed models and time series clustering to analyze trends by state. The association of comorbidities and other variables with the risks were studied by fitting a spatial panel data linear model (splm). As results, we observed that on average the greatest risks were reached by July, and that highest risks were observed in some states, Baja California Norte, Chiapas, and Sonora; interestingly, some densely populated states, as Mexico City, had lower values. Different trends by state were observed, and a four-order polynomial, including fixed and random effects, was necessary to model them. The most general structure is one in which the risks increase and then decrease and was observed in states belonging to two clusters; however, there is a cluster corresponding to states with a retarded increase, and another in which increasing risks through time were observed. A cyclic behavior in terms of states having a second increasing trend was observed. Finally, according to the splm, percentage of men, being in the group of 50 years and over, chronic kidney disease failure, cardiovascular disease, asthma, and hypertension were positively associated with the case-fatality risks. This analysis may provide valuable insight into COVID-19 dynamics in future outbreaks, as well as the determinants of these trends at a state level; and, by combining spatial and temporal information, provide a better understanding of COVID-19 case-fatality.

## 1. Introduction

Coronavirus disease (COVID-19), which is caused by the SARS-CoV-2 virus (Severe Acute Respiratory Syndrome Coronavirus type 2), was first reported in Wuhan, China, on December 31th, 2019 (World Health Organization, 2020), and on March 11th, 2020, the World Health Organization (WHO) declared a pandemic state. On that date, the number of cases of COVID-19 outside China increased 13-fold and the number of affected countries had tripled. There were more than 118,000 cases in 114 countries and 4,291 people lost their lives. Of these cases, more than 90 percent were in just four countries, and two of those -China and the Republic of Korea-had significantly declining epidemics, whereas 81 countries had not reported any cases, and 57 countries had reported 10 or less cases (World Health Organization, 2020, March 11th). On April 20th, the COVID-19 disease spread rapidly and on October 19th, 2020, more than 40.6 million cases of the disease had been reported in 220 countries and territories in the world, being the five countries with the highest number of infected the United States, India, Brazil, Russia, and France, with more than 1.1 million deaths, whereas the United States, Brazil, India, Mexico, and the United Kingdom were the five countries with the highest number of deaths.

An increased risk in suffering severe COVID-19 occurs if the infected person belongs to the elderly group and with underlying health conditions, such as cardiovascular disease, chronic kidney disease, chronic respiratory disease, chronic liver disease, diabetes, cancer, HIV/AIDS, tuberculosis (active), chronic neurological disorders, sickle cell disorders, smoking tobacco use, obesity (BMI ≥40), and hypertension (Pan American Health Organization, 2020, July 29th) according to guidelines published by the WHO, CDC, and Public Health England (PHE).

In Mexico, the first reported case of COVID-19 was detected on February 28^th^. By April 30th, 63 days after this first diagnosis, the number of patients exponentially increased, reaching a total of 17,799 confirmed cases and 1,732 deceased. As of June 26^th^ 2021, Mexico had confirmed 2,503,408 cases and 232,521 deaths (INFOBAE, 2021). Moreover, the National Institute of Statistics (INEGI) reported in 2021 that from January to August 2020 the deaths from COVID-19 occupied the second cause of death in Mexico with almost 108,658 cases below cardiovascular diseases (141,873 cases), and above diabetes mellitus (99,733 cases). In contrast, in 2019, 88.8% (663,902) of deaths in Mexico were due to illnesses and health-related problems and the three main causes of death for both men and women were heart disease (156,041, 23.5%), diabetes mellitus (104,354, 15.7%), and malignant tumors (88,680, 13.4%) (INEGI, 2020, October 29th). Additionally, according to the latest National Health and Nutrition Survey (ENSANUT), Mexico ranks as one of the countries with the most number of obese people in the world, being such disease a public health problem (Barquera, et al., 2020). Comorbidities associated with COVID-19 severe cases in Mexico coincide with some of these causes of death, these comorbidities have been found to be ageing, hypertension, obesity, diabetes, and smoking, as well as chronic kidney, cardiovascular problems, COPD, immunosuppression, and asthma (Gobierno de la Ciudad de México, 2020).

Spatial epidemiology analyses describe geographic variations in a disease with respect to demographic, environmental, behavioral, socioeconomic, genetic, and infectious risk factors, and they have proven to be quite useful to study the spread of infectious diseases analyses (Elliott & Wartenberg, 2004). In this sense, they could help us to highlight areas and communities which could be hotspots or coldspots, pinpoint cases, measure risks and map transmission in a cost-saving way, and to improve the targeting of limited resources (Tatem, 2018). Several examples of spatial epidemiology analysis have demonstrated its importance such as for Tuberculosis (Shaweno et al., 2018), Ebola (Mizumoto et al., 2019), or Malaria (Mercado et al., 2019), or through mathematical models based on the behavior of gases to understand the COVID-19 propagation in Mexico City (Salcido, 2021), to mention a few examples. Particularly, concerning COVID-19, several studies in France (Levratto et al. 2020) and Italy (Bourdin et al., 2021) indicated that the demographic composition (Sannigrahi et al., 2020), such as population density, of a country influences the rate of fatalities due to COVID-19. Additionally, in a recent publication with data from twelve European countries, it was shown that the number of medical practitioners and hospital beds and the level of social trust are correlated with low COVID-19 death rates (Amdaoud et al., 2021). Moreover, a spatial analysis performed in the USA demonstrated that COVID-19 could move from less vulnerable counties to more vulnerable counties, and back again over time (Neelon et al., 2021). Even so, it was demonstrated that demographics explain the spatial heterogeneity in COVID-19 testing and infection rates in New York City neighborhoods (Schmitt-Grohé et al., 2020). In a recent published study by Argoty-Pantoja et al., the authors found that COVID-19 adversely affects the indigenous population, particularly patients who received initial outpatient care (Argoty-Pandoja et al., 2021).

Here, we performed a spatial epidemiological study of case-fatality risks due to the COVID-19 in Mexico. Spatio-temporal analyses are performed concerning risks by state in Mexico mainly from April to September, including descriptive analyses through mapping and time series representations. Additionally, we analyze trends through time by state by using linear mixed models and time series clustering. Finally, the association of the main comorbidities with the COVID-19 death risks are studied by fitting spatial panel data linear models.

## 2. Material and methods

### 2.1. Data sources

We used open-source data of suspected COVID-19 cases collected by the General Directorate of Epidemiology (DGE), of the Ministry of Health of Mexico (*Dirección General de Vigilancia Epidemiológica, Secretaría de Salud* (Secretaría de Salud, 2020)). The dataset was updated till September 30th, 2020. Two datasets were created, one weekly and other monthly. The starting date for the weekly database was April 1th, 2020, date on which all states had at least one death, which was used for most analyses, except for the spatial linear model in which April 15^th^, 2020, was used since till that date there were positive cases in each combination of time and state, whereas April 1st, 2020, was used for the monthly database. Weekly deaths were obtained since January 1st, 2020, to generate an animation as explained below. We used the date when the patient entered the respective hospital unit, and in the case of deaths, the date when the patient died.

For the assembly of the weekly and monthly datasets, we considered the following: only the cases with positive results for SARS-CoV-2 were selected and the data were grouped by state and by date, weekly and monthly, respectively. For each level of grouping, we obtained the number of positive cases, number of deaths, number of men, number of people by age group (groups constructed every 10 years, but as discussed below, we finally used the age group of individuals 50 years and over), and number of pregnant women. In addition, for each of the comorbidities presented in the DGE dataset, the number of patients who had a positive diagnosis for each of the following comorbidities (in alphabetical order) were counted: asthma, cardiovascular diseases, chronic kidney failure, diabetes, COPD, hypertension, immunosuppression, pneumonia, and obesity. As the response variable, we used the case-fatality risk, calculated as the number of deaths over the number of positive cases grouped by entity and date. The remaining variables were used as explanatory in the spatial panel data linear model.

### 2.2. Analyses

#### 2.2.1. Descriptive space-time analyses

We obtained weekly and monthly case-fatality risks associated with each time and state. Monthly risks were taken since April 2020, and weekly risks since January 2020 and also since April 2020. The corresponding maps were derived and a space-time plot (Hovmöller diagram), that shows the risks in a space-time cross section, was obtained. An animation (gif) associated with maps corresponding to weekly risks since January was obtained, and the time series associated with weekly risks by state since April 2020 were plotted. R packages *spacetime* and *magick* were used for the maps and gif analysis.

#### 2.2.2. Analyses of the trends structure by state

To determine trends for each state, linear mixed models (LMM) (Verbeke & Molenberghs, 2000) were fitted for the weekly data since April 15th, 2020; thus, associating random effects with each state. First, we fitted a LMM including constant and linear terms, both as fixed and random effects. A map of linear trends by state was obtained since all random and fixed effects were significant according to a Likelihood Ratio test (LRT; comparing with a model without random effects) and t-tests; respectively. To have a better understanding of the type of trend, polynomic trends were added. From the linear trend model, random effects corresponding to polynomials of different degree were sequentially added. For instance, a second-order random effect was added to the linear model, which was significant according to an LRT, and thus added to the model. This new model was compared with one adding a third-order random effect, and so on, until a random effect associated with certain order was not significant. According to this process, a four-order polynomial was used, including each term as random and fixed, being all significant according to an LRT (comparing with a model without random effects) and t-tests, respectively. The original and predicted time series by state were plotted. R package *nlme, sp, ggplot2*, and *lattice* were used to the trends structure by state analysis.

#### 2.2.3. Spatial panel linear models

Spatial panel linear models (splm), which are a variant of a panel data regression including spatial effects, were fitted, e.g. (Pebesma, 2012), (Millo & Piras, 2012), (Wikle, Zammit-Mangion, & Cressie, 2019). A spatial weights matrix is required for this process, thus, we obtained Queen contiguity weights. Hence, a squared matrix of dimension 32 (the number of states in Mexico) was obtained with all entries equal to zero or one, the latter indicating that two states are neighbors. The weights are calculated by integrating this matrix in a row-standardized form. Variants of splm can be fitted according to the presence or not of different terms: a space lagged effect associated with the response, a space lagged effect associated with the error term, and the presence or not of random or fixed effects proper of panel data analyses. The risks were used as response variable and the remaining variables as explanatory. Each observation corresponds to a time-state combination. All explanatory variables were used as percentages with respect to the number of infected people. Age was firstly obtained according to 10-year age groups but was finally used as the percentage of individuals aged 50 years and over, since this aggregated age group was more associated with the risks and all the age-groups forming it are positively associated with them.

The response variable was transformed into different scales, but we finally used a logit transformation since the normality assumption was better satisfied in this scale. We added a small constant (summing around 0.5 deaths by observation), chosen in each model as that value improving the normality assumption, since in the logit transformation logarithms are used and there are zero values in some observations. Association between explanatory variables and between them with the response was analyzed by using association measures (Pearson, Spearman, and Kendall). We fitted univariable models to determine which variables were significantly associated with the risks, eliminating from the multivariable model those that were not significant. We analyzed the linearity assumption of each input with the response by obtaining scatter plots including a smoothed LOESS (locally estimated scatterplot smoothing) curve. Since we observed that few important outliers in some variables were present, we replaced them for more appropriate values. The splm with the variables obtained after this process was fitted and model assumptions were assessed. We determined whether the space-lagged effect and random effect were significant, jointly and conditionally, by using Baltagi, Song and Koh LM-H one-sided joint test and conditional LM tests, and whether random or fixed effects were preferable by using a Hausman test for spatial models. R packages *splm, spdp, rgdal, ggplot2*, and *corrplot* were used for the splm analysis.

#### 2.2.4. Time series clustering

Time series clustering is a method used to create groups or clusters of dynamic data, e.g. (Sardá-Espinosa, 2017) and (Sardá-Espinosa, 2019). The members of the same cluster are similar with each other, but dissimilar from objects in a different cluster. In analysis of clustering for time series, it should be considered the measure of similarity or distance, the function of prototype or centroid extraction (a time series derived from the data which is representative of a cluster), the clustering algorithm, and the cluster evaluation. The most common clustering procedures are hierarchical and partitional methods. The clustering of time-series proposed by (Aghabozorgi, Shirkhorshidi, & Wah, 2015) may be shape-based, feature-based, or model-based. In the shape-based method is common to utilize the dynamic time warping (DTW) distance as dissimilarity measure. In this paper, we used a partitional clustering, a DTW distance, a k-medoids method (Partition Around Medoids or PAM), which means that a time series in the data set is used as a representative of a cluster, and an average linkage. All the time series were standardized. To evaluate the clusters, we variated the number of clusters (3 to 9) and compared them using the silhouette as an internal evaluation measure. R package *dtwclust* was used to cluster the data.

## 3. Results

### 3.1. Results of descriptive space-time analyses

The gif presented in **Supplementary Material** corresponds to the evolution through time of case-fatality risks in Mexico by state and by week from January 1st to September 30th, 2020. During the early spread of COVID-19 in Mexico, high case-fatality risk levels were present in northwest states (mainly Baja California and Chihuahua) and in center states (Mexico, Morelos, Hidalgo, and Guerrero). Moreover, on average the highest mortality rates were observed in July, followed by June and August. On average, higher case-fatality risks were observed mainly in Baja California Norte (state in the border with United States of America), Chiapas (state in the border with Guatemala), and Sinaloa; followed by Campeche, Hidalgo, Mexico, and Morelos (the last three in the center of Mexico, and neighbors of Mexico City), and other states showing lower risks as Baja California Sur, Tamaulipas, and Yucatán.

This also can be identified in the Hovmöller diagram (**Figure 1a)**), and from it and **Figure 1b)**, which shows trends by state along time, we observed that there were states with greater case-fatality risks at the beginning and then a decrease, as in Durango and Tabasco, other states in which the peak of the risks was earlier, and a few states as Chiapas in which this peak was reached much later. Monthly case-fatality risks by state are presented in **Figure 1c)**.

**Figure 1:**
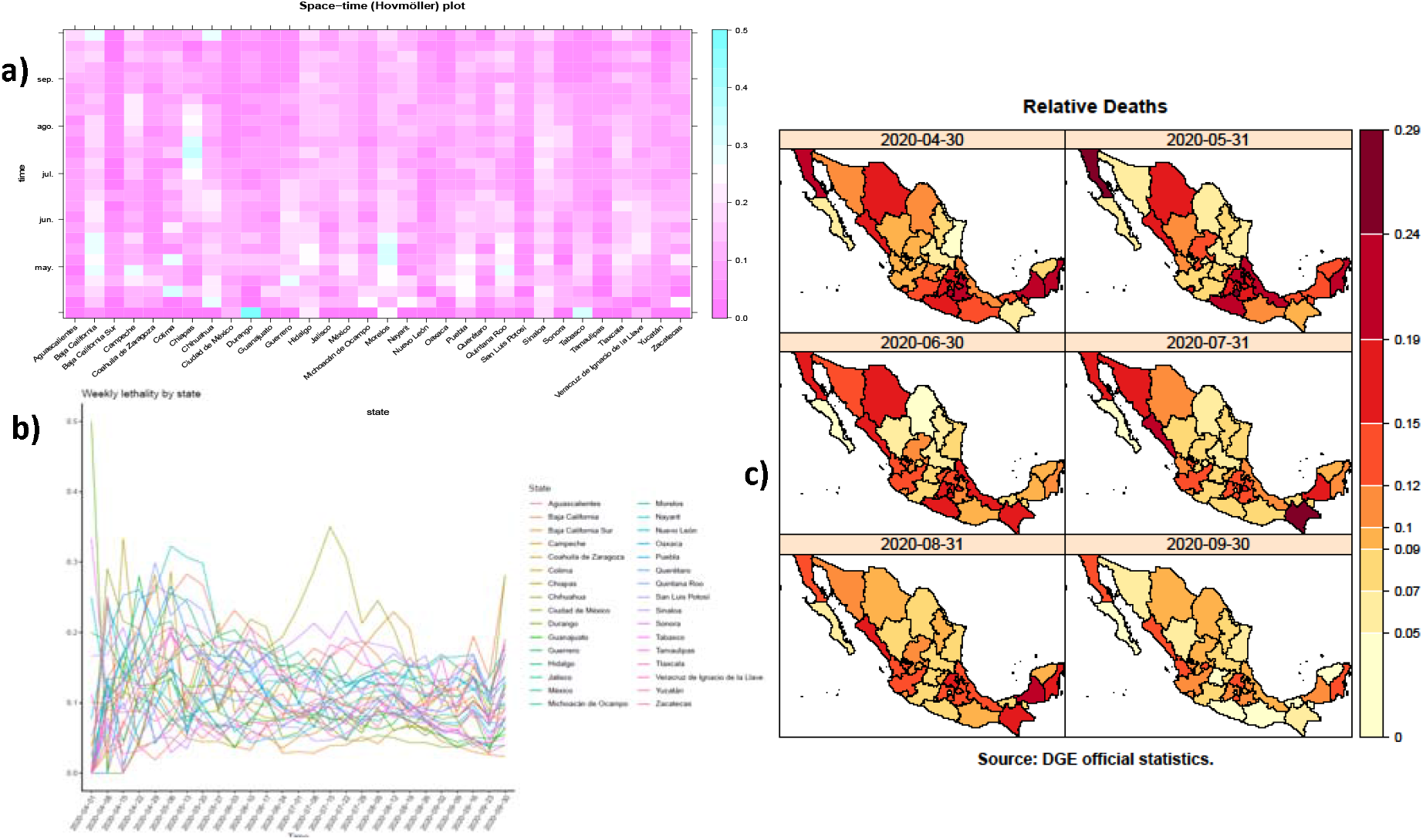
Representations of COV ID-19 case-fatality risks in Mexico by state from April 1^st^ to September 30^th^, 2020: a) Space time (Hovmöller) plot representing weekly risks in a space-time cross section (a color gradient is presented in which purple represents less risks and blue greater risks), b) Weekly time series (trends) associated with each state, and c) Monthly risks.

### 3.2. Results of analyses of trends structure by state

Considering case-fatality risks as response and time as explanatory variables, the fixed effects for both constant and linear terms were significant (t= 29.912, p-value<0.001; and t=-3.420, p-value<0.001; respectively). The corresponding random effects were jointly significant when compared with the model including only the fixed effects (LR=295.056, p-value<0.001). A map corresponding to this linear trend (including both fixed and random effects) is shown in **Figure 2a)**. States have different case-fatality risks trends. We could group the states by trend levels as follows: Aguascalientes has the highest (positive, around 0.0035), followed by Chiapas, Jalisco, San Luis Potosí and Tamaulipas (in orange, around 0.0015), and Campeche and Nayarit (in salmon, around 0.001); others states were around zero, for instance Coahuila de Zaragoza, Nuevo León, and Querétaro (in pink, around 0.0005), Sonora (in magenta, around 0.0), and Baja California Sur, Tlaxcala, Veracruz de Ignacio de la Llave, Yucatán and Zacatecas (in purple, around −0.0005). Finally, other states have the lowest negative trend levels, for instance, Baja California Norte, Colima, Guanajuato, Michoacán de Ocampo, Oaxaca, Sinaloa, and Mexico (in blue violet, around −0.0015), followed by Hidalgo, Mexico City, and Quintana Roo (in blue, around −0.002), Chihuahua and Durango (in dark blue, around −0.0025), and Puebla (in navy, around −0.0035), and finally Guerrero, Morelos, and Tabasco (in dark navy, around - 0.0045).

**Figure 2:**
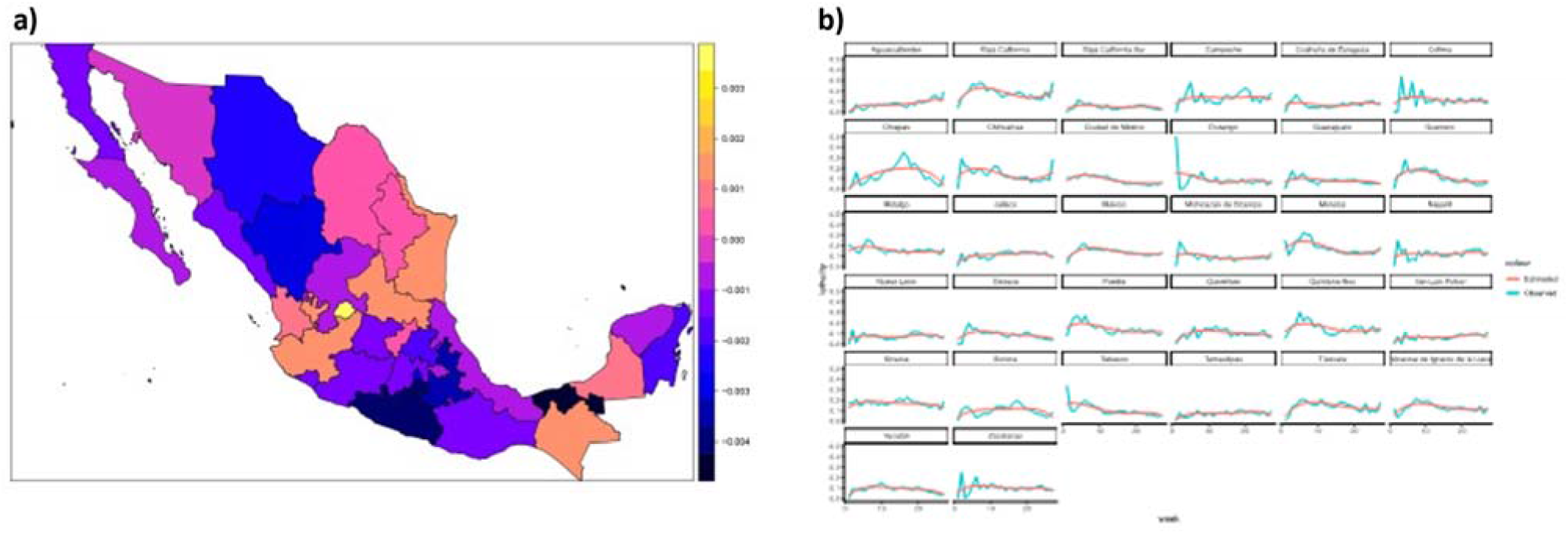
Representations of trends estimated through Linear Mixed Models (LMM): a) Map of the linear trend by state obtained using an LMM with fixed and random effects for constant and linear terms and b) Observed and predicted case-fatality risks obtained from a LMM with fixed and random effects corresponding to a four-order polynomial.

We added to the model including only linear effects some polynomial trends as random factors. The quadratic trend was significant (LR=57.908, p-value<0.001) and thus added to the model. In the new model, the cubic trend was significant (LR=72.855, p-value<0.001) and added, obtaining another model in which the quartic trend was also significant (LR=16.044, p-value=0.007), and thus added to the model. However, considering the last model, a five-order trend was not significant (LR=7.517, p-value=0.2756), thus, the model corresponded to a four-order polynomial. The corresponding fixed effects were all included and were significant (t tests with p-value<0.001 for each fixed effect); and in this, our final model, all random effects were jointly significant (LR=413.095, p-value>0.001). In **Figure 2 b)** the original and predicted time series by state are plotted. The graphs show the estimated values, i.e. the marginal mean profiles, obtained from the LMM, in which a four-order polynomial with intercept including both fixed and random effects for state was fitted. Notice that some trends in average increase as in Aguascalientes, Jalisco, and San Luis Potosí. Other states have mixed trend patterns: first increasing, then decreasing, and at the end of the study increasing as in Baja California Norte, Chiapas, Chihuahua, Coahuila de Zaragoza, Morelos, and Nuevo León. Meanwhile, in other states there are constant trends as in Baja California Sur, Campeche, Colima, Durango, Guanajuato, Hidalgo, Michoacán de Ocampo, Nayarit, Quintana Roo, Sinaloa, Sonora, Mexico, Tamaulipas, and Zacatecas. Finally, we observed decreasing trends in Guerrero, Mexico City, Oaxaca, Puebla, Querétaro, Tabasco, Tlaxcala, Veracruz de Ignacio de la Llave, and Yucatán. Notice that the highest levels of the average trends are in Baja California Norte, Chiapas, Morelos, and Sinaloa, presenting the highest case-fatality risks.

### 3.3. Spatial panel linear models

In **Figure 3**, we show the association (Pearson correlation) between the relative number of men, people in the age group of 50 years and over, pregnant women, and the comorbidities using in the logit transformation with a constant term of 0.5, whereas in the Supplementary material (**Figure S1**) we present a similar figure including the response and 10-year age groups, in which it can be seen that it was convenient to aggregate age. Similar results were obtained using other association measures. We observed high association (greater than 0.5) between people aged >50 years and diabetes, hypertension, pneumonia, and the highest association between comorbidities, corresponded to diabetes with hypertension and diabetes with pneumonia. Pneumonia had the highest association with the logits; however, since an association in both directions between this variable and the response is suspected, and even there is some confusion between them, we decided to eliminate it.

**Figure 3.**
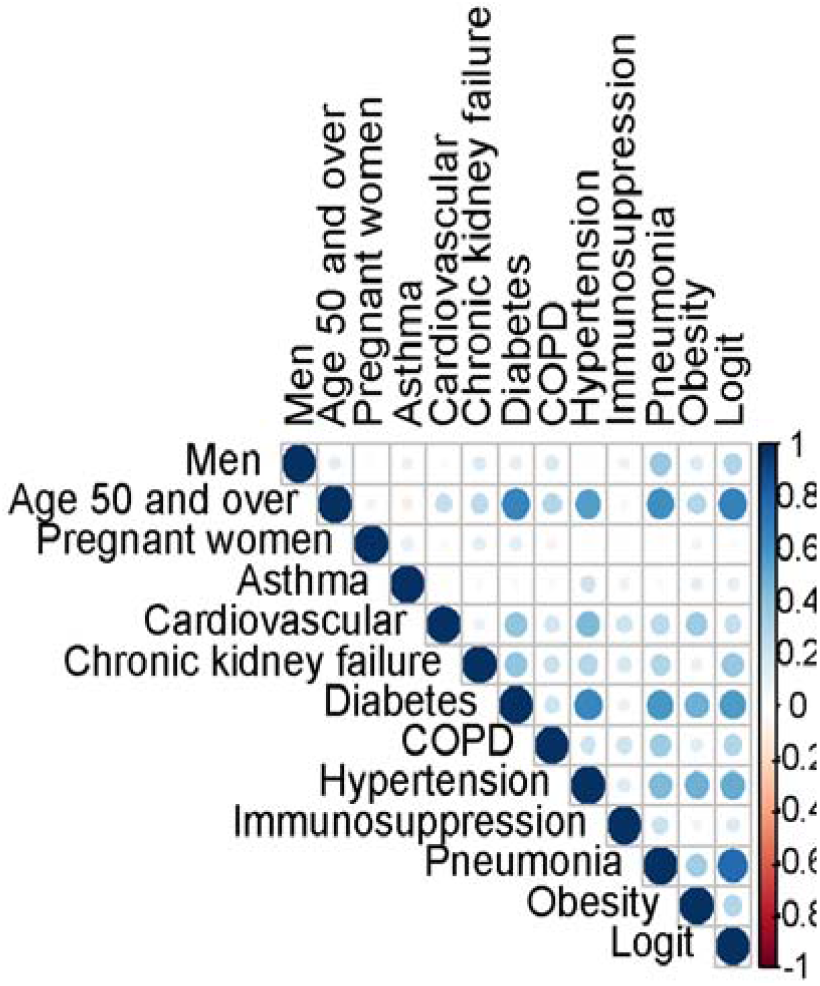
Association between inputs and between them and the output, the logit transformation of the case fatality risks: men (%), age group 50 years and over, pregnant women, and prevalence associated with the comorbidities.

To fit the spatial lineal model, first we obtained the Queen contiguity matrix indicating the neighbors of each state (**Table S1**). Then the corresponding weights matrix was built. Significant variables in the univariable spatial linear models were number of men (%), people aged >50 years (%), and in terms of comorbidities prevalence of asthma, cardiovascular diseases, diabetes, COPD, hypertension, immunosuppression, pneumonia, obesity, and chronic kidney failure (**Table 1**). The percentage of pregnant women was not significant and thus not included in the multivariable model. From the significant variables, we examined the linearity assumption and observed that two variables (asthma and cardiovascular diseases) had one outlier each one, corresponding to earlier observations in time in which few positive cases were present; thus, we replaced them for the corresponding mean **(Figure S2, Supplementary Material**).

**Table 1.**
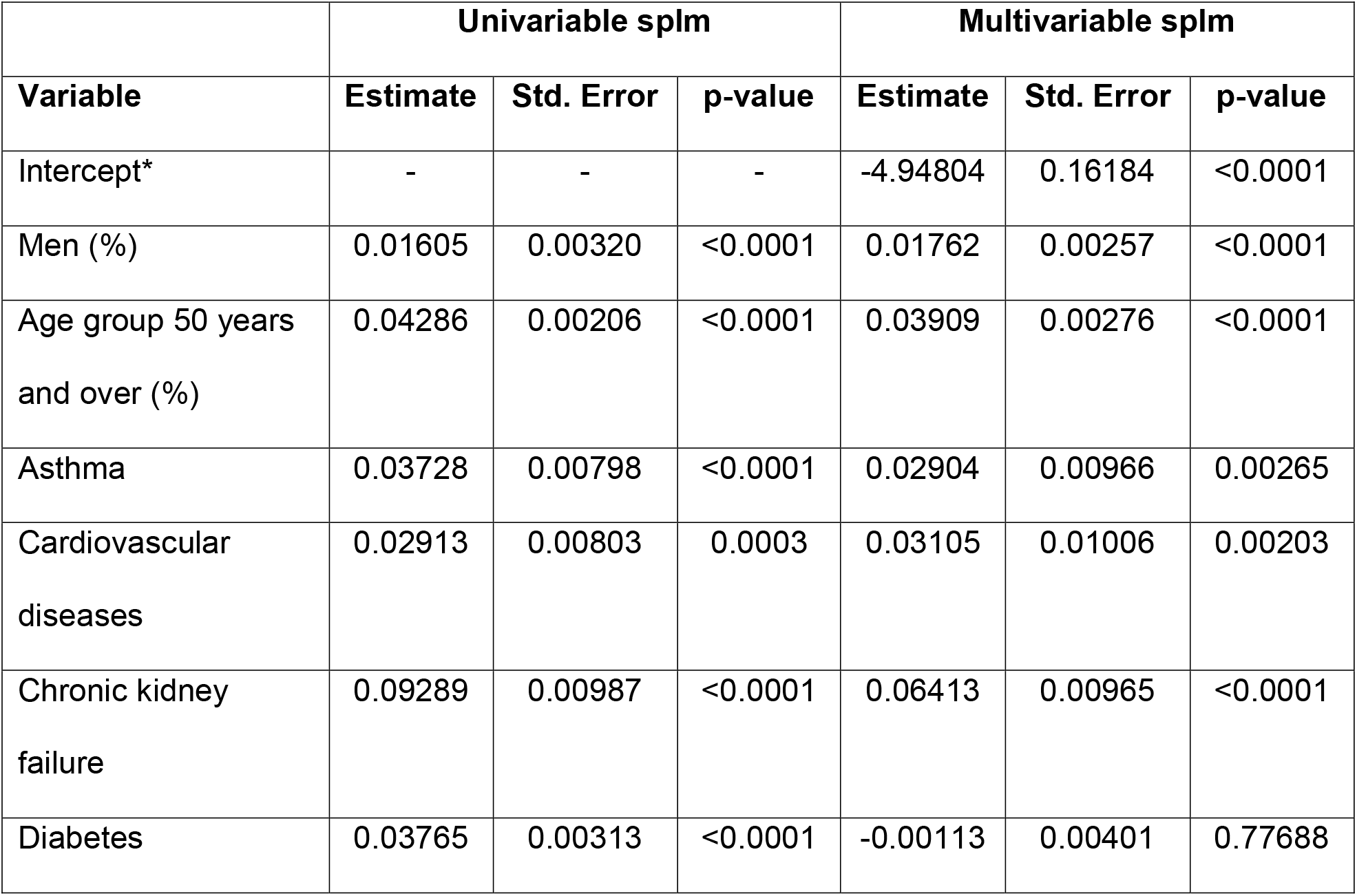
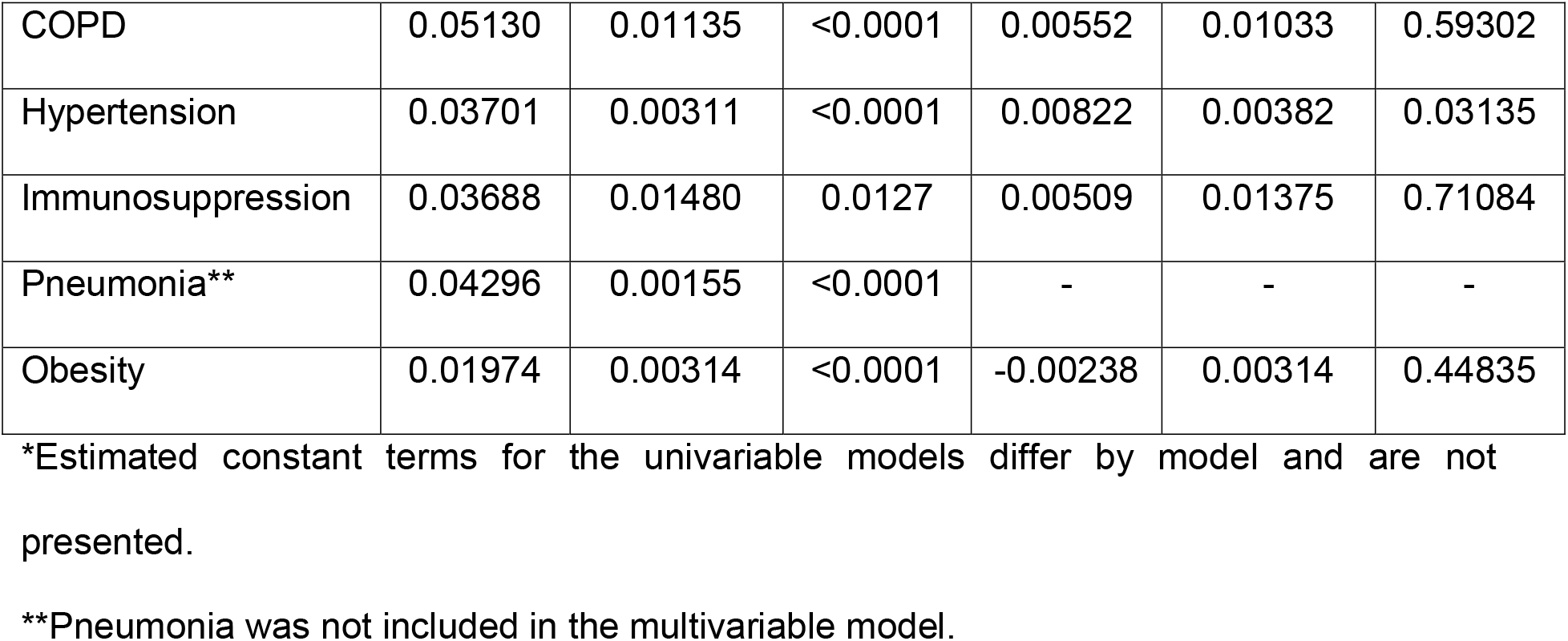
Univariable and multivariable spatial panel linear models between selected inputs (percentage of men, people aged 50 years and over (%), and prevalence of asthma, cardiovascular diseases, chronic kidney failure, diabetes, COPD, hypertension, immunosuppression, and obesity) and case-fatality risks.

|  | Univariable splm |  |  | Multivariable splm |  |  |
| --- | --- | --- | --- | --- | --- | --- |
| Variable | Estimate | Std. Error | p-value | Estimate | Std. Error | p-value |
| Intercept* | - | - | - | -4.94804 | 0.16184 | <0.0001 |
| Men (%) | 0.01605 | 0.00320 | <0.0001 | 0.01762 | 0.00257 | <0.0001 |
| Age group 50 years and over (%) | 0.04286 | 0.00206 | <0.0001 | 0.03909 | 0.00276 | <0.0001 |
| Asthma | 0.03728 | 0.00798 | <0.0001 | 0.02904 | 0.00966 | 0.00265 |
| Cardiovascular diseases | 0.02913 | 0.00803 | 0.0003 | 0.03105 | 0.01006 | 0.00203 |
| Chronic kidney failure | 0.09289 | 0.00987 | <0.0001 | 0.06413 | 0.00965 | <0.0001 |
| Diabetes | 0.03765 | 0.00313 | <0.0001 | -0.00113 | 0.00401 | 0.77688 |
| COPD | 0.05130 | 0.01135 | <0.0001 | 0.00552 | 0.01033 | 0.59302 |
| Hypertension | 0.03701 | 0.00311 | <0.0001 | 0.00822 | 0.00382 | 0.03135 |
| Immunosuppression | 0.03688 | 0.01480 | 0.0127 | 0.00509 | 0.01375 | 0.71084 |
| Pneumonia** | 0.04296 | 0.00155 | <0.0001 | - | - | - |
| Obesity | 0.01974 | 0.00314 | <0.0001 | -0.00238 | 0.00314 | 0.44835 |

\*Estimated constant terms for the univariable models differ by model and are not presented.
\*\*Pneumonia was not included in the multivariable model.

Since the correlation matrix did not reveal high correlation between the inputs (**Figure 3**), and thus multicollinearity was not suspected, we used in the multivariable model all inputs, except for pneumonia and pregnancy as discussed before. Hence, the multivariable spatial linear model included percentage of men, people aged 50 years and over (%), and prevalence of asthma, cardiovascular diseases, chronic kidney failure, diabetes, COPD, hypertension, immunosuppression, and obesity (**Table 1**). We determined that the random effects (mu) and spatial effects associated with the response (lambda) were jointly significant (LM-H=566.71, p-value<0.001) and significant when conditioned to one another (LM*-mu=25.477, p-value<0.001 and LM*-lambda = 6.471, p-value<0.001; for the random effects and spatially lagged effect, respectively), that random effects were necessary (Hausman Chisq=0.800, p-value=0.999), and that the spatial lagged effect for the error term (rho) was also significant (t=6.187, p-value<0.0001). Some interaction effects were considered, but since they caused multicollinearity problems, they were omitted from the model. The residuals obtained from the LMM by using all covariates do not show neither problems of lack of normality nor lack of constant variance (**Figure S3** in Supplementary Material). Results associated with the multivariable model are given in **Table 1**. All the variables, except diabetes, immunosuppression, COPD, and obesity, were significantly associated with the risks. Results show that the percentage of men and being in the group of 50 years and over are positively associated with the case-fatality risks (OR= 1.01778, p-value<0.001; OR=1.0399, p-value<0.001, respectively). The comorbidities significantly associated with higher fatality risks were chronic kidney disease failure (OR=1.0662, p-value<0.001), cardiovascular disease (OR=1.0315, p-value=0.002), asthma (OR=1.0294, p-value=0.003), and hypertension (OR=1.008, p-value=0.030).

### 3.4. Time series clustering

According to the silhouette criterion, four groups of time series were necessary. The prototypes (centroids) of each cluster correspond to the states of Baja California Sur (cluster 1), San Luis Potosí (cluster 2), Mexico City (cluster 3), and Sonora (cluster 4). The names of each state by group are displayed in **Figure 4**.

**Figure 4.**
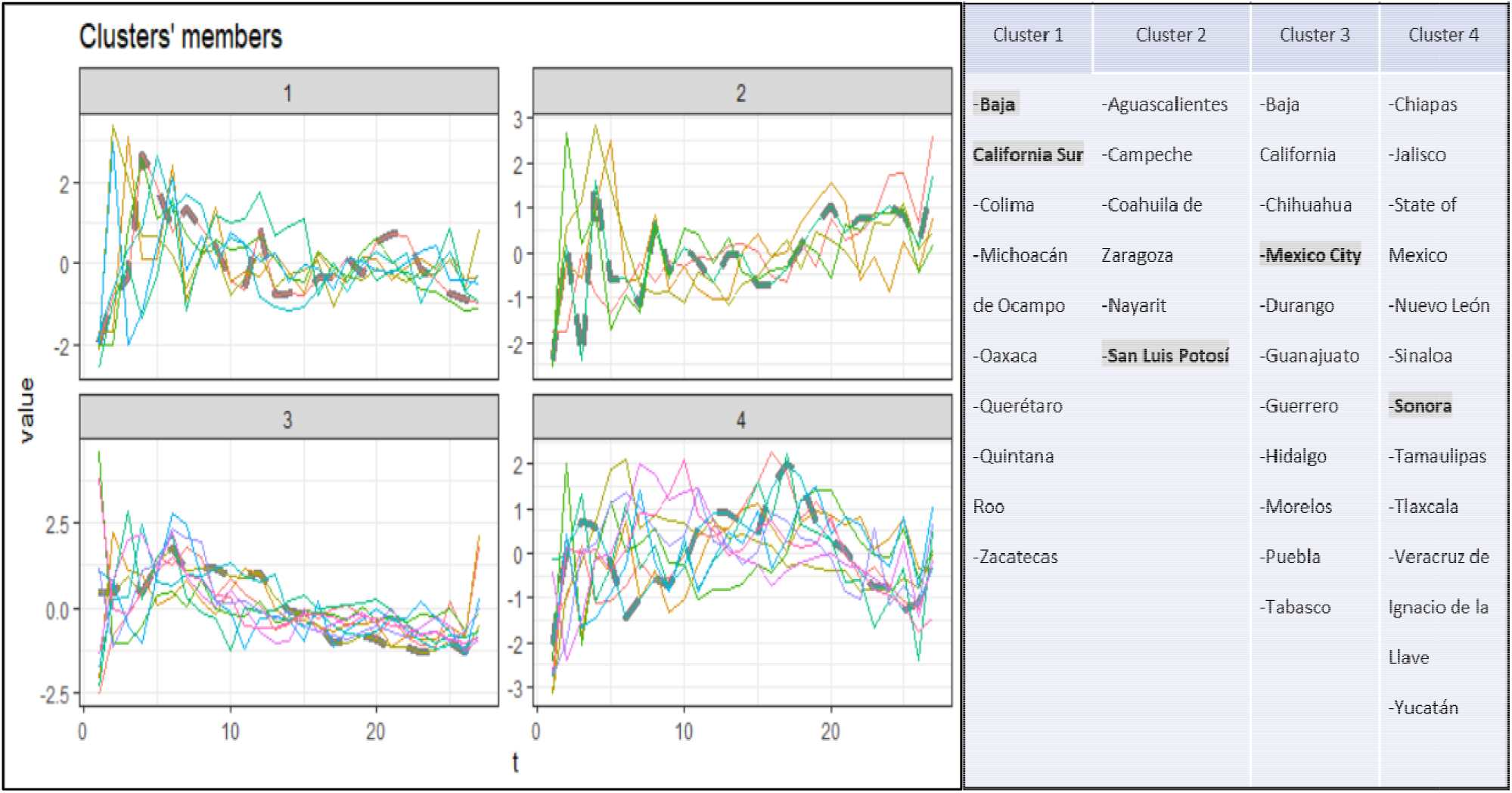
Groups of COVID-19 case-fatality risks trends (time series clustering) by state. They are obtained using a partitional clustering method, a DTW (dynamic time warping) distance, k-medoids, and average linkage. Centroids are shown as gray dashed lines and correspond to the states of Baja California Sur (cluster 1, with seven elements), San Luis Potosí (cluster 2, with five elements), Mexico City (cluster 3, with ten elements), and Sonora (cluster 4, with ten elements). Cluster members are also shown, the centroids of each cluster are in bold.

The groups are obtained according to the risk trends by state, we observed four available trajectories of the standardized risks along time (**Figure 4**). Group 1 includes seven states and displays a high increasing trend at the beginning and then a slow decreasing trend, having at the middle of the analyzed time of observation in average a constant trend. Note that in general these states have kept their constant levels, as Colima or Querétaro. Group 2 includes five states and shows an increasing trend. Note that these states are those who at the beginning maintained low risks, but along time these were growing as in Aguascalientes or San Luis Potosí. Group 3 includes ten states, showing high fatality risks from the beginning then a slow decrease and at the end these remained more or less constant. Notice that these states had the highest fatality risks from the beginning, as Baja California, Mexico City, or Morelos. Finally, Group 4 includes ten states and shows a high increase of the risks at the beginning, then a constant trend at the middle, and finally a decreasing trend at the end of the time of observation. The trends in these states are more dissimilar than in other groups, having a wider range of variation, for instance, this group includes states such as Chiapas which had high risks from the beginning and Yucatán that had had low values all the time.

## 4. Discussion and Conclusions

Here, we applied different spatio-temporal analyses concerning COVID-19 case-fatality risks. First, descriptive analyses through maps and animations allowed us to understand how COVID-19 case-fatality risks evolved through time, in what can be considered as the first wave of COVID-19 in Mexico and the beginning of the next wave. We observed how the risks differentially increased in all the territory, though this increase seemed to be concentrated firstly in the Northwest, Center, and Yucatán peninsula. Afterwards, in other states, as Jalisco or Chiapas, the risks increased obtaining the highest values, together with those corresponding to the same zones in which the risks increased from the beginning. Of notice is how Chiapas has some of the highest values, which may be related with the presence of indigenous groups as some studies through survival analyses indicate (Argoty-Pantoja et al. 2021). In this sense, it has even been shown that the association with COVID-19 cases spatially varies according to a disadvantage measure and indigenous composition (Huyser at al. 2021). Interestingly, Mexico City, despite having the greatest number of deaths and positive cases, did not reach the same high risks as in other states, which may be attributable to higher detection of mild and asymptomatic cases compared to the rest of the country. We also observed that at the end of the period of analysis, the risks increased again, interestingly; once again a greater increase was observed in some states in the North and Yucatan Peninsula, which might suggest a kind of cyclic spatial behavior. However, care should be taken with the results associated with the first weeks, since few COVID-19 cases were identified; and thus, the risks seem to be larger than those of future weeks. This same issue was observed when the spatial linear model was fitted in terms of other variables, and this was the reason why percentages associated with two variables had to be replaced by their mean. Thus, we think risks assessments are more reliable after April or May.

The spatial linear model had a good fit and allowed us to use the longitudinal (through time) and spatial information (by state), improving the precision of the estimated associations when compared with a cross-sectional analysis. The dynamic of the risks varies through time, thus results of different cross-sectional analysis could provide different pictures. To improve the estimation process, we used a small constant term in each model, which allowed us to improve the normality assumption. Additionally, by considering that both time and space generate correlation between observations, we improve the estimations. For the multivariable model, we eliminated pneumonia as an input since pneumonia is not necessarily a precondition, as other comorbidities, and can be caused by the COVID-19 infection. Thus, we think that an analysis of the direction of the association between COVID-19 and pneumonia could be relevant in future research. According to the splm results, the variables that were significantly positively associated with the case-fatality risks were percentage of men, percentage of individuals in the 50 years and over age group, and prevalence of chronic kidney disease failure, cardiovascular diseases, and asthma. Notably, comorbidities associated with the distribution of COVID-19 in Mexico had previously been identified. An older population structure and a high burden of cardio-metabolic comorbidities predisposes individuals to have more severe disease, thus increasing overall mortality trends (Bello-Chavolla et al., 2020 and 2021). Northern states in Mexico are characterized by having higher rates of chronic cardio-metabolic conditions, which may explain some of the higher case-fatality rates observed in this study. Whether the distribution of metabolic burden may have the primary influencer of case-fatality or whether this could be attributable to intrinsic deficiencies in healthcare systems which worsen access and quality of medical care remains as an opportunity for future research. When analyzing clusters of the time series associated with the case-fatality risks, we used both partitional and hierarchical clustering methods. We presented here the partitional method since for hierarchical clustering, we obtained a cluster formed by only one state. However, the structure of the time series pertaining to each cluster were similar independently of the method used. It is interesting to see how there were different pattern behaviors in how the risks evolved through time, the most common pattern, as seen in Group 1 and 3, corresponded to states with an increase and then a decrease of the case-fatality risks through time, and afterwards a constant trend. However, some states showed a retarded increase, and in some states, Group 2, even an increase through time. Probably these differences are related with mobility and how in some states the disease was propagated much earlier than in others. In this sense, we can clearly see how in the north frontier, particularly around the Baja California Peninsula, Yucatan, and near Mexico City, the increase in the risks was earlier and remained high through all the time window of analysis. This can be explained by considering that in these places there is more external mobility, and around Mexico City, a higher population density, economic activity, and internal mobility, which may be the clue for an earlier increase in the risks (Ramírez-Aldana et al, 2021). Similarly, social vulnerability could also be behind the difference in trends between states, as has been shown when comparing death rates in counties in the United States (Neelon et al. 2021) with high and low vulnerability and in marginalized municipalities within Mexico City (Antonio-Villa et al. 2021). The difference between death rates by country in Europe has also shown this differential of fatality through time and by county (Amdaoud et al., 2021), showing also that there are spatial clusters of regions that evolve through time, though it is difficult to define all the factors behind this behavior.

The trends could also be examined through the fitted linear mixed models, in which we used random and fixed effects and identified that a four-order polynomial was necessary to properly model the trends associated with the risks. The estimated trends allowed us to identify that the most general structure is one in which the risks increase and then decrease; though, as we saw in the clusters, there are some states in which this type of trend is not present. When considering only linear trends, we observed than in most states this was decreasing or around zero, which makes sense considering that by analyzing the first wave, in most states there was at the beginning an increasing trend and then decreasing; however, once again there are some states with some increasing trend, which once again can be related with the difference in the propagation of the disease through all the territory. It is important to understand the different trends by state, to identify what differs in them. Perhaps, identifying those states with a more successful decrease in the risks, we could understand how people was treated there, to apply this successful scheme in other states or future waves.

It is important to notice that the results we obtained are aggregated by state and time. In this sense, the results, particularly in terms of the spatial lineal model, cannot be inferred to an individual level (Pearce, 2000). For instance, we know that a higher percentage of individuals aged ≥50 years is associated with greater risks, but we cannot derive from this result that being an individual ≥50 years and over is associated with higher risks. Thus, all conclusions in terms of the model must be taken with care. However, the aggregated analyses allowed us to better understand trends and clustering and association with the risks at a state level, considering more information than in any cross-sectional or retrospective analysis.

In terms of the information, we must consider that the number of COVID-19 cases is probably larger, and that the number of deaths could vary as well. Thus, the risks are only an indicative of the real values. In this sense, in our analyses we used the information from January to September and not afterwards, in part, to consider that the definition of a positive case was changed using other criteria from this date. If further spatio-temporal analyses including dates after and before October were performed, this matter should be considered, and the risks should be properly adjusted. For future research, a similar analysis could be performed at a municipality level (spatial unit contained in a state). In this sense, recently there have been efforts to provide aggregated data at this level using the same data set we used for our analysis, including additional variables (Mas, 2021). However, a spatio-temporal analysis of COVID-19 case-fatality in Mexico as the one we present here has not been previously performed.

In conclusion, we demonstrated a heterogeneous profile in the distribution of case-fatality risks across Mexico during the first wave of COVID-19 during 2020. State profiles point at spatially defined units which may have influenced how COVID-19 mortality occurred during this first wave and may provide valuable insight into COVID-19 dynamics in future outbreaks, as well as some determinants of these trends at a state level. By combining spatial and temporal information, a more in-depth understanding of COVID-19 case-fatality may inform public policies for regional pandemic management.

## Supporting information

Animation

Weekly data

Weekly data from January

Monthly data

## Data Availability

Data available as supplementary material

## Supplementary Material

Supplementary material is available at the journal webpage. A gif with maps corresponding to weekly case-fatality risks since January 2020 is included. The reader is referred to the online Supplementary Material for more Figures and Tables.

## Notes

### Competing Interest Statement

The authors have declared no competing interest.

### Funding Statement

No external funding was received

### Author Declarations

All the information in which this study is based on is freely available and does not require any ethics commitee aproval

