## Supplementary tables and Figure for "A spatio-temporal study of state-wide case-fatality risks during the first wave of the COVID-19 pandemic in Mexico"

**Title:**

**1. Results of descriptive space-time analyses**

A gif with maps corresponding to weekly risks since January 2020 is available at the journal webpage.

**2. Results of spatial panel linear models**

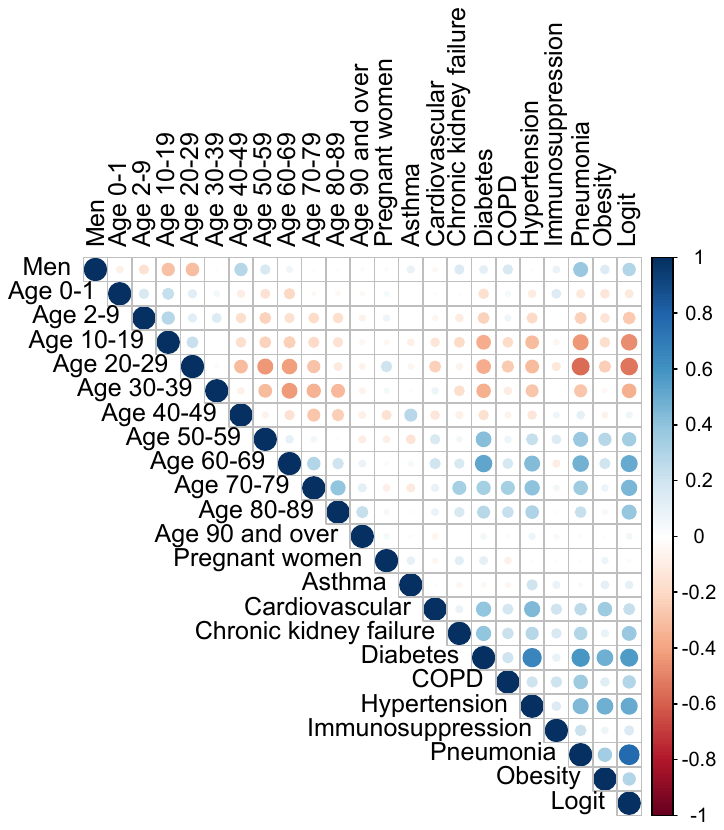

Figure S1. Association between inputs and between them and the output (transformed case-fatality risks): men (%), age group (%), pregnant women (%), and prevalence of the comorbidities.

Table S1. Neighbors for the 32 states of Mexico according to a Queen contiguity matrix.

| **State ID** | **State Name** | **Neighbors** |
| --- | --- | --- |
| 1 | Aguascalientes | 14, 32. |
| 2 | Baja California Norte | 3, 26. |
| 3 | Baja California Sur | 2. |
| 4 | Campeche | 27, 31. |
| 5 | Coahuila de Zaragoza | 8, 10, 19, 32. |
| 6 | Colima | 14, 16. |
| 7 | Chiapas | 20, 27, 30. |
| 8 | Chihuahua | 5, 10, 25, 26. |
| 9 | Mexico City | 15, 17. |
| 10 | Durango | 5, 8, 18, 25, 32. |
| 11 | Guanajuato | 14, 16, 22, 24, 32. |
| 12 | Guerrero | 15, 16, 17, 20, 21. |
| 13 | Hidalgo | 15, 21, 22, 24, 29, 30. |
| 14 | Jalisco | 1, 6 ,11, 16, 18, 32. |
| 15 | State of Mexico | 9, 12, 13, 16, 17, 21, 22, 29. |
| 16 | Michoacán de Ocampo | 6, 11, 12, 14, 15, 22. |
| 17 | Morelos | 9, 12, 15, 21. |
| 18 | Nayarit | 10, 14, 25, 32. |
| 19 | Nuevo León | 5, 24, 28, 32. |
| 20 | Oaxaca | 7, 12, 21, 30. |
| 21 | Puebla | 12, 13, 15, 17, 20, 29, 30. |
| 22 | Querétaro | 11, 13, 15, 16, 24. |
| 23 | Quintana Roo | 31. |
| 24 | San Luis Potosí | 11, 13, 19, 22, 28, 30, 32. |
| 25 | Sinaloa | 8, 10, 18, 26. |
| 26 | Sonora | 2, 8, 25. |
| 27 | Tabasco | 4, 7, 30. |
| 28 | Tamaulipas | 19, 24, 30. |
| 29 | Tlaxcala | 13, 15, 21. |
| 30 | Veracruz de Ignacio de la Llave | 7, 13, 20, 21, 24, 27, 28. |
| 31 | Yucatán | 4, 23. |
| 32 | Zacatecas | 1, 5, 10, 11, 14, 18, 19, 24. |

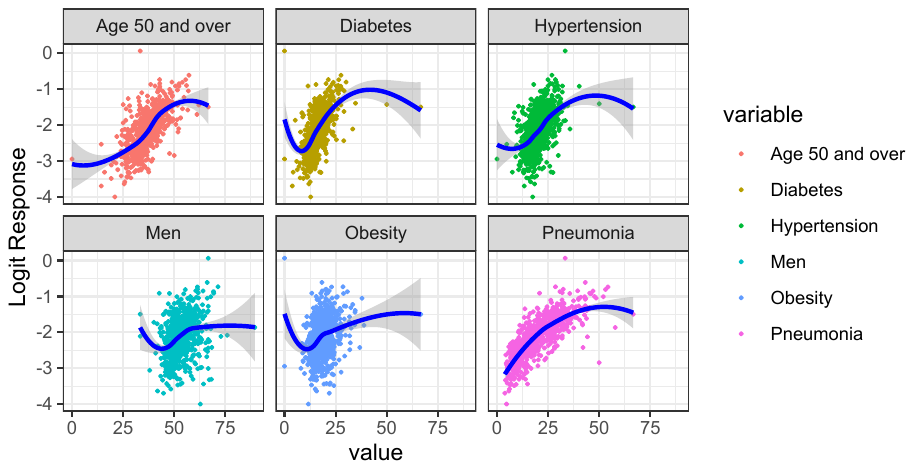

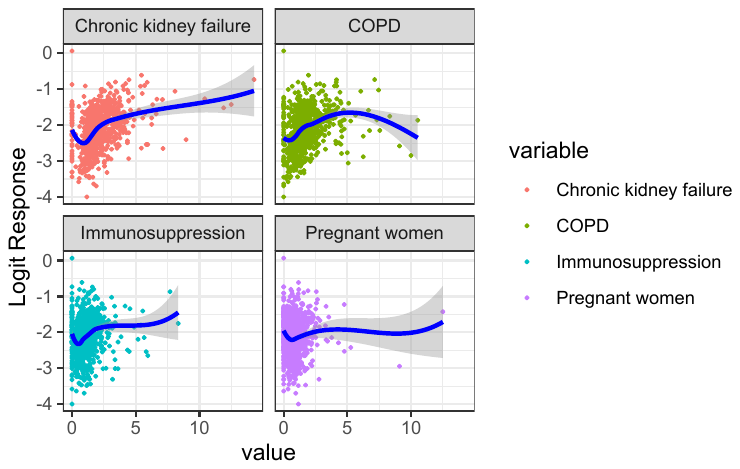

Figure S2 a) Results of the univariate analysis using LOESS for each variable.

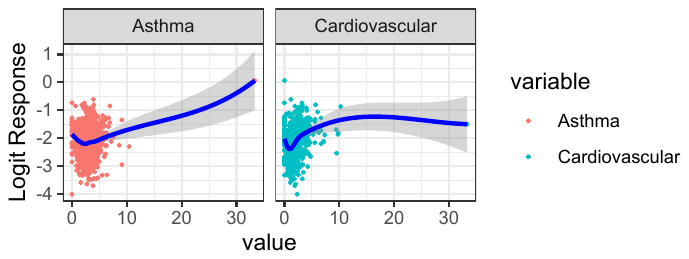

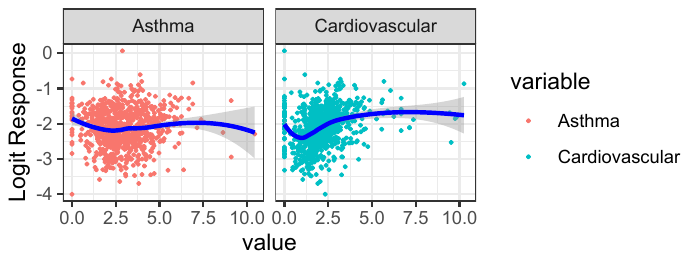

Figure S2 b) Results of the univariate analysis using LOESS for asthma and cardiovascular diseases, for which there are outliers, which were replaced by the mean.

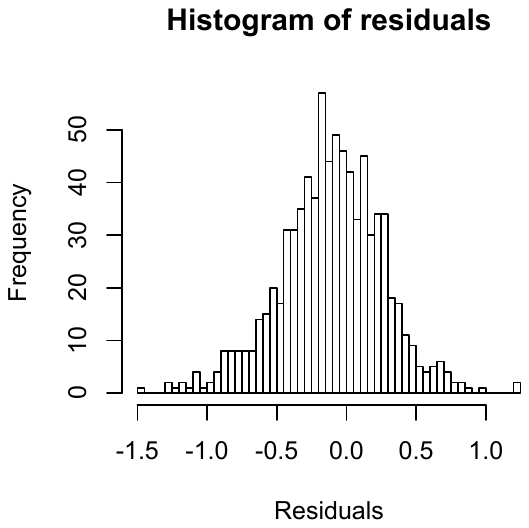

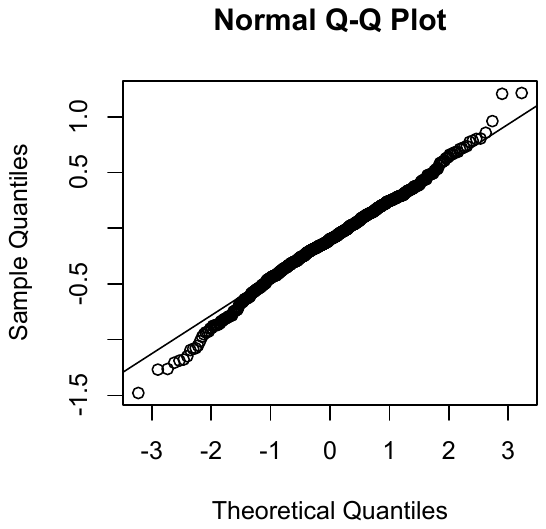

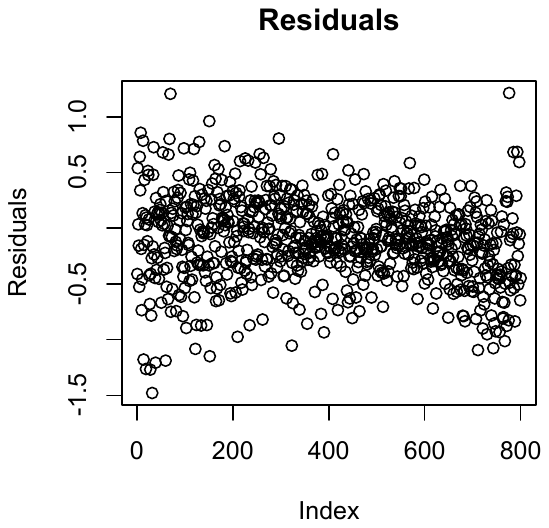

Figure S3. Residuals of the fitted spatial linear model including as inputs percentage of men, people aged 50 years and over (%), prevalence of asthma, cardiovascular diseases, chronic kidney failure, diabetes, COPD, hypertension, immunosuppression, and obesity.
