## Supplementary figures and images for "A spatio-temporal study of state-wide case-fatality risks during the first wave of the COVID-19 pandemic in Mexico"

### Animation

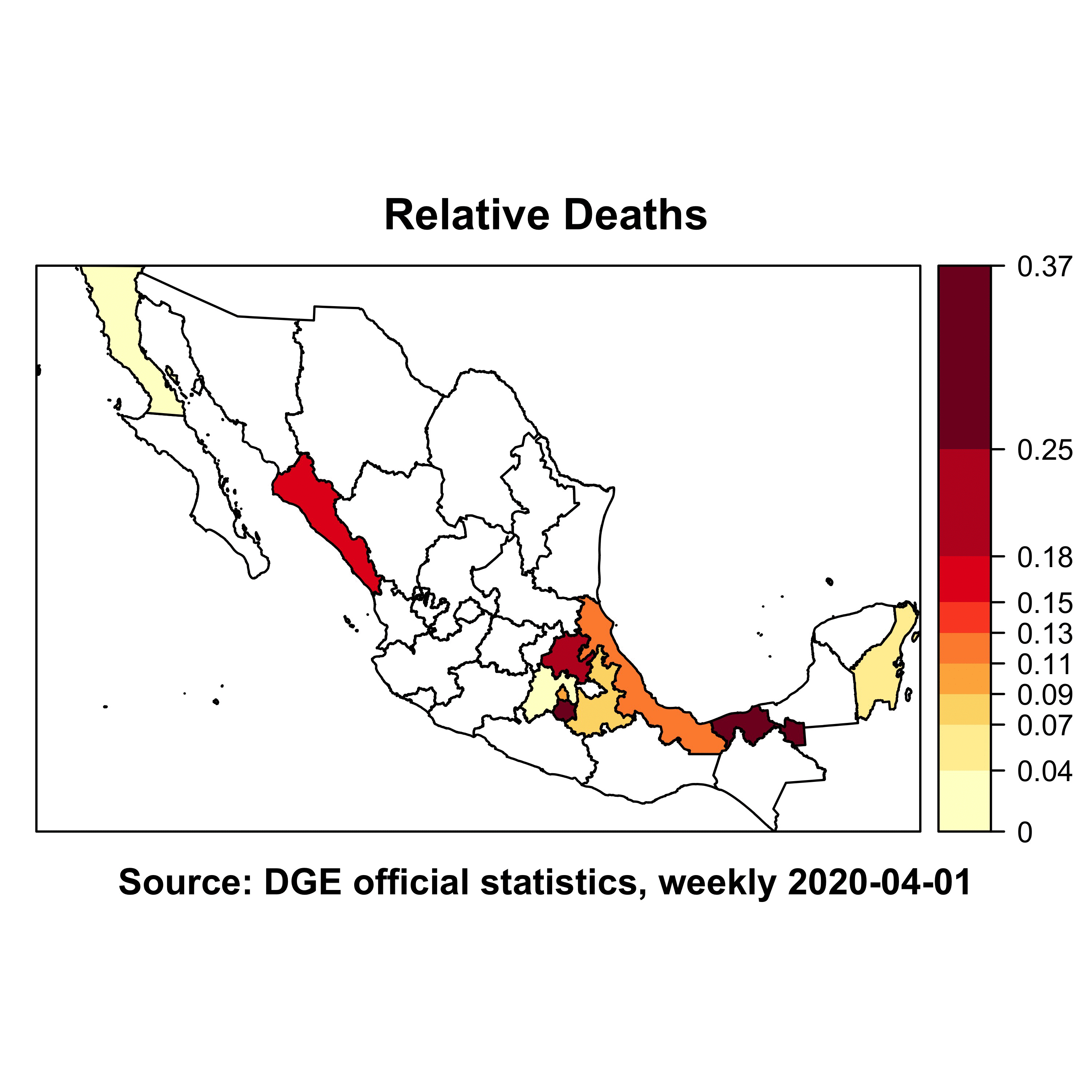
